# Computational model of neuronal recruitment during ICMS for restoring somatosensation in the human somatosensory cortex

**DOI:** 10.1101/2023.06.22.23291761

**Authors:** Pawel Kudela, Matthew S Fifer, Luke E Osborn, David P McMullen, Pablo A Celnik, Gabriela L Cantarero, Francesco V Tenore, William S Anderson

## Abstract

**Objective:** Intuitively providing touch feedback from artificial hands to users with sensory loss remains a challenge. Although localized fingertip sensations can be evoked via intracortical microstimulation (ICMS), feedback is generally optimized using psychometric tasks rather than mimicking the cortical response to touch.

**Approach:** We created an anatomically-informed and participant-specific model of the human somatosensory cortex (S1) region with an implanted microelectrode array (MEA). We performed simultaneous stimulation-and-recording from the study participant S1 region to characterize cortical responses elicited by single ICMS pulses. Pulses were delivered to a set of pre-selected electrodes mapped to tactile receptive fields. We next performed a 2D (i.e., in the plane of the MEA probe tips) current source density (CSD) analysis of recorded cortical responses to inform cortical network model parameters on how ICMS activates neurons and lateral synaptic connections in the area of the S1 sampled by MEA electrodes. Using information from planar CSD profiles obtained from ground truth data, we reconstructed lateral connections in the S1 model needed to produce the desired responses to single ICMS pulses. The effect of multiple ICMS was then simulated in the biologically realistic cortical model and the results were validated against ground truth cortical responses from the study participant.

**Main results:** A high-resolution cortical network model, calibrated to produce the known cortical responses to single ICMS pulses delivered to individual electrodes, predicted with a reasonable accuracy the cortical response to ICMS pulses delivered simultaneously to multiple electrodes.

**Significance:** These preliminary results suggest that high-resolution biologically realistic cortical network models can potentially be reliable predictors of cortical response to a given pattern of ICMS presentations and therefore useful in designing biomimetic stimulation patterns.

## Introduction

Brain-machine interface (BMI) approaches have enabled individuals with paralysis to control the movement of cursors and robotic arms (Aflalo *et al* 2015, Bouton *et al* 2016, Collinger *et al* 2013, Hochberg *et al* 2012, Wodlinger *et al* 2015). Increasingly, intracortical microstimulation (ICMS) has been employed as a strategy for providing somatosensory feedback to BMI users (Flesher *et al* 2016, Armenta Salas *et al* 2018, Flesher *et al* 2021). Feedback strategies fall along a gradient in their biomimicry from learned artificial mappings (O’Doherty *et al* 2011, Dadarlat *et al* 2015), to psychometrically-derived encoding models (Flesher *et al* 2021, Osborn et al 2021, Tabot *et al* 2013), to a growing contingent of encoding strategies designed to replicate cortical responses (Bensmaia and Miller 2014, Kumaravelu *et al* 2020, Saal *et al* 2017). Encoding strategies that derive parameters from theoretical models and/or neurophysiological responses benefit from a greatly reduced parameter search space and hold promise for deriving more intuitive and robust tactile percepts.

Using a biomimetic stimulation approach creates several challenges, however—the effect of ICMS applied to a given volume of tissue is difficult to predict and largely unknown. Theoretical studies suggest that, due to nonlinear interactions between stimulation-induced electric fields and neural and synaptic activations, a superposition-principle of individual ICMS is insufficient (Hokanson *et al* 2018). Previous theoretical studies, however, either do not explicitly model the effects of the electric field on axonal processes of simulated neurons (i.e., current injected to a single compartment model (Berger *et al* 2012)) or lack the complexity to simulate neuronal responses resulting from cortical network architectures (Hokanson *et al* 2018). Other factors such as electrode design, location, orientation, and gyral geometry also may also significantly impact model findings (Kudela and Anderson 2015, Kudela and Anderson 2021, Thielscher *et al* 2011).

We have implemented a high-resolution computational model of neuronal recruitment during ICMS in the human somatosensory cortex. Our objective is to build a forward computational model for predicting cortical responses in the somatosensory cortex to the delivered ICMS pulses. We hypothesize that the computational models with an appropriate level of complexity can aid in determining the sequence, amplitude, and frequency of ICMS pulses needed to replay spatiotemporal patterns in the somatosensory cortex associated with tactile percepts.

We sought to determine whether a cortical network model with reconstructed functional connectivity required to produce the desired cortical responses to ICMS pulses delivered to individual MEA electrodes could predict the outcome of ICMS pulses delivered to multiple MEA electrodes simultaneously. This question has been addressed previously only in theoretical studies but has not been quantitatively investigated *in situ*. Here we simulated the effect of multichannel ICMS in the biologically realistic cortical model and validated the results against ground truth neural recordings from a human subject. We first created an anatomically-informed and participant-specific model of a segment of the human somatosensory cortex with an implanted microelectrode array (MEA). We recorded cortical responses evoked by stimulation within the somatosensory cortex of the study participant elicited by single ICMS pulses delivered to a set of pre-selected electrodes that mapped to tactile receptive fields. We next analyzed local field potentials of the recorded cortical responses which contains information about how the dendrites process synaptic inputs. We used a 2D current source density (CSD) analysis to quantify these responses in the plane of MEA probe tips and inform the cortical network model parameters on how ICMS presentations activate neurons and lateral synaptic connections in the area of the somatosensory cortex sampled by MEA electrodes. Using information from planar CSD profiles obtained from ground truth data, we reconstructed lateral connections in the somatosensory cortex model needed to produce the desired responses to ICMS pulses. We demonstrated that simulated cortical responses were largely consistent with the recorded cortical responses in the study participant. Finally, we simulated cortical responses to ICMS pulses delivered to all pre-selected electrodes simultaneously, and compared the model-predicted cortical responses to the recorded cortical responses in the somatosensory cortex of the study participant. The model with the reconstructed lateral connections predicted cortical responses to ICMS delivered to all electrodes well on a fine timescale as assessed by comparing the model-derived planar CSD profiles to the recorded (ground truth) responses. These results suggest that high-resolution biologically realistic cortical network models can potentially be reliable predictors of cortical response to a given pattern of ICMS presentations and therefore useful in designing biomimetic stimulation patterns. The optimization of biomimetic ICMS patterns for restoring somatosensation can be possibly performed by selecting the ICMS patterns such that model-based responses (2D CSD) match the measured responses (2D CSD templates) during natural tactile sensations.

## Methods

### Finite Element Model of the somatosensory cortex

Folded surface and heterogeneous conductivity of the cerebral cortex require numerical methods to reveal the electric field potential generated by a stimulating electrode in extracellular space. To quantify the distribution of the electric potentials generated by ICMS delivered to an individual MEA electrode, we created a high-resolution, anatomically-informed, and study participant-specific finite element model (FEM) of a segment of the somatosensory (S1) cortex with an implanted MEA. All finite element analyses were performed in COMSOL Multiphysics 5.3 (Burlington, MA). The outer (pial) surface of the postcentral gyrus (Fig 1a) was obtained from the tissue segmented 7T-MRI image of the subject’s left hemisphere using FreeSurfer image analysis suite v7.1.0 (Dale *et al* 1999, Fischl and Dale 2000). The 12 mm-long section of the gyrus containing S1 contralateral to the stimulated hand was extracted (region of interest) Fig 1.c The gyral folding in the region of S1 overlapping with the area of MEA implantation (putative Brodmann’s area 1) was reconstructed (Fig 1e). During the reconstruction process, the volume of the extracted gyrus in the region of interest was sliced by 12 parallel cross-section planes separated 1 mm apart along the gyral crest line. A quintic Bezier parametric curve was fitted to each pial contour line for each gyral cross-section plane independently. The 3D reconstruction of the gray matter (GM) superficial boundary (i.e., pial surface, Fig 1d) was performed by plotting the interpolated 3D-surface based on the obtained 72-coordinate point array (12 planes x 6 Bezier’s control points). The representation of the GM superficial border in analytical form allowed for the creation of the high-resolution finite element model of the putative Brodmann’s area 1 which was then coupled to compartmental representations of cortical neurons for analyzing neuronal activation by ICMS. The reconstructed region of interest (Fig 1f) was segmented into the GM (3 mm thick, conductivity σ=0.26 S m^-1^) and white matter (WM) (σ=0.126 S m^-1^) segments and the space above the gyrus was filled with cerebrospinal fluid (CSF) (σ=1.65 S m^-1^). The conductivities of the GM, WM, and CSF were modeled as isotropic and σ values were obtained from the literature (Koessler *et al* 2017, Nicholson 1965, Baumann et al 1997). The array of 32 MEA contacts was reconstructed and included in the model (Fig 1d). The exact horizontal and vertical positions of individual MEA probe tips with respect to the cortical surface cannot be determined because the size of a probe tip is beyond the resolution of MRI/CT. The post**-**implant CT scan coregistered with the pre-implant MRI were used to determine the MEA horizontal position for the FEM model. In the vertical direction, MEA probe tips were positioned 1 mm (MEA probe shank length) beneath the reconstructed pial surface. Electrode contacts were modeled as cones, 50 μm high with a diameter of 60 μm at the base tapering to a tip (after the manufacturer’s specifications; Blackrock Microsystems, Salt Lake City, Utah). Spacing between electrodes was 400 μm. The entire model (stacked segments of WF/GM/CSF) had dimension 12 x 12 x 6 mm and contained 1,063,367 tetrahedral FEM elements (Fig 1f).

**Fig 1.**
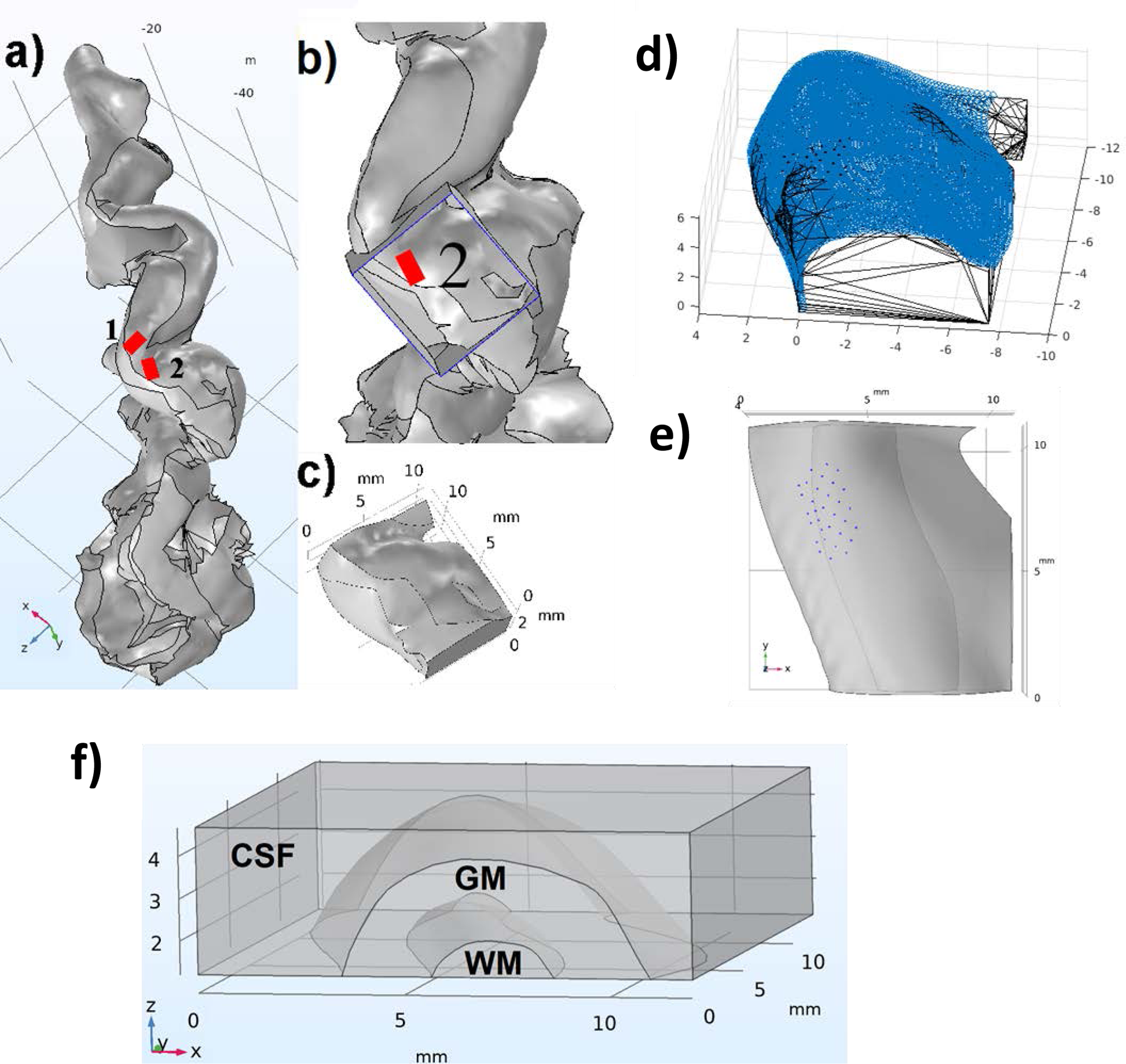
a) The outer (pial) surface of the study participant’s postcentral gyrus. A raw gyral geometry (NIFTI/STL) from MRI segmentation was converted to a surface object with 1 and 2 indicating sensory areas implanted with MEAs (marked in red). b) The block (violet) indicates the section of interest with the implanted MEA #2 in area 1 of S1. c) The extracted section of interest from the block in b). d) The 3D parametric surface (blue) fitted to gyral anatomy in the area 1 of S1. e) The reconstructed section of interest of S1 (top view) along with the reconstructed MEA contact positions (black dots). f) The volume conductor model for the FEA was segmented into gray matter (GM), white matter (WM), and cerebrospinal fluid (CSF).

Monopolar stimulations were used to collect ground truth data for the model calibration. Surface boundary current sources (von Neumann boundary conditions) were specified to control current values at the targeted electrodes. The current density at the electrode was calculated by dividing the amplitude of the applied ICMS current (85 μA) by the total exposed surface area of the electrode (0.0055 mm^2^). To mimic the effects of monopolar stimulation (return electrode “at infinity”) ground nodes were placed around the exterior boundary of the finite element mesh (i.e. current sinking to the outer boundaries of the model) (Butson and McIntyre 2005). We verified that the total integrated current density over the outer boundaries of the FEM was equal to the total integrated current density over the exposed surface area of the electrode (inner boundary of the model). Capacitive and inductive currents were not included in the FEM, which implies that the medium is purely resistive and is referred to as the quasi-static model.

The network model and the FEM shared a common coordinate system. The FEM delivered electric potential values inside the modeled cortical volume at the time of electrical stimulations for each compartment of each simulated pyramidal neuron. Compartment coordinates were exported from the GENESIS simulator before network simulations. During network simulations, the values of the external electric potential from the FEM analyses for the obtained coordinates were used to calculate the equivalent current injections for all compartments of simulated neurons as described in Rattay F 1999 and Warman *et al* 1992. Amplitudes of equivalent current injections were calculated for each compartment and these currents were injected into compartments at the time course of stimulation. The membrane potential of the axonal compartments were checked against a potential threshold of 0 mV to determine which axonal compartment was activated first after the ICMS presentation. Additional details can be found in our previous work (Kudela and Anderson 2015, Kudela and Anderson 2021).

### Computational model: Cortical network

All network simulations were performed in parallel GENESIS 2.3 (http://genesis-sim.org/GENESIS) (Bower and Beeman 1998, Bower and Beeman 2007). The pyramidal cell model of a regular-spiking neuron was adopted from our previous modeling studies of the effects of subdural cortical stimulations. Details of the neuron morphologies have been described earlier and can be found in (Kudela and Anderson 2015, Kudela and Anderson 2021). In short, the key element of the network simulations was a 156-compartment model of a layer 2/3 neocortical pyramidal neuron (Fig 2a). For the network simulations, we arranged 173,056 (416 x 416) pyramidal neurons to form a supergranular cortical layer 2/3 inside the cortical gyrus.

**Fig 2.**
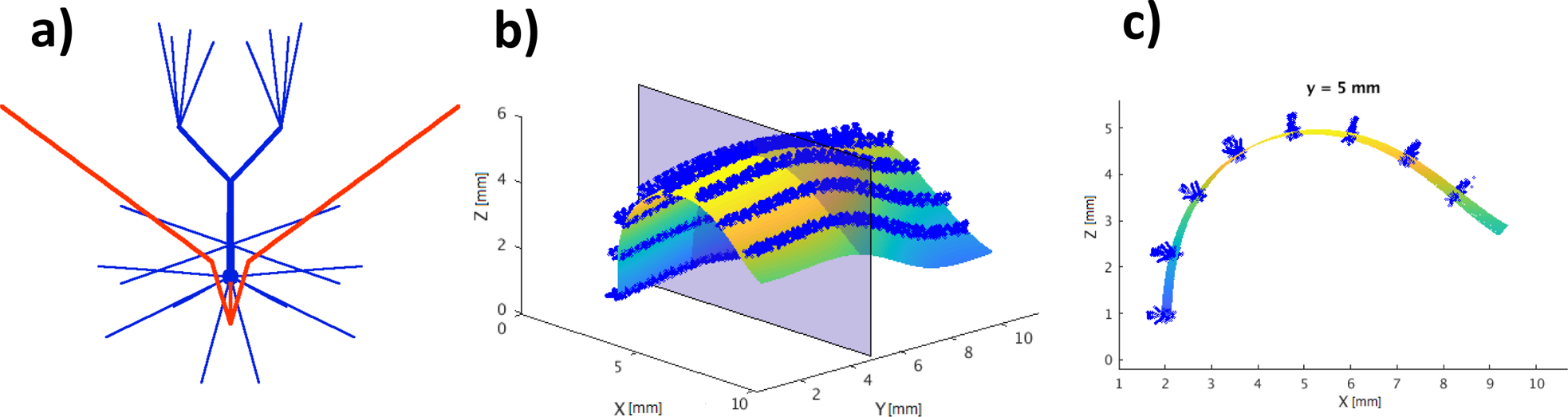
a) The model of the pyramidal neuron with the dendritic (blue) and axonal (red) compartments. B) The 3D parametric surface was used for the cortical layer 2/3 reconstruction in the somatosensory cortical network model. The blue brush-like strips illustrate the positions of pyramidal neurons (dendritic compartments) distributed along the Y axis on the surface for the selected eight X coordinates (note that only a few rows are shown for better visibility). c) The parametric curve (XZ planes shown in b)) from the parametric surface fitted to the gyral anatomy. Curves were used to govern the positions and orientations of the pyramidal neurons (only dendritic compartments shown) across cortical layers in a 3D mode of S1. The total number of neurons in the model was 173056 (416×416).

The 3D arrangement of pyramidal neurons in the cortical layer followed the subject-specific gyral folding pattern in the region of interest (Fib 2b). The apical-basal axis of each neuron was oriented perpendicularly, while axon branches projected tangentially to the pial surface of the gyrus (Fig 2c). We used a parametric representation of the reconstructed pial surface (gray matter superficial boundary) to populate pyramidal neurons at equal distances from the pial surface and to create a 0.8 mm thick supragranular cortical layer 2/3 beneath the gyral surface. Each neuron was rotated by a randomized angle (0° - 360°) about its apical-basal axis. The final 3D coordinates of each neuron inside the created cortical layer were also randomized (within a given 5% range). This allowed for the arrangement of neurons inside a cortical layer in an anatomically realistic fashion and analogous to the cortical arrangement of neurons in gyri. In horizontal directions (i.e. parallel to the gyral surface) distance between next-nearest neurons was 49 µm. The arrangement of neurons in the horizontal direction provided a spatial scale, making a total area of a simulated cortical patch equal to approximately 141.6 mm^2^ with 1221 neurons/mm^2^. Within the simulated cortical layer, pyramidal neurons were connected with AMPA chemical synapses. These connections represent the horizontal (lateral) components of cortical connectivity targeting apical dendrites of the pyramidal neurons. The probability of the connection between pyramidal neurons was 5% up to 400 µm (measured from the axon terminal). Each pyramidal neuron contacted approximately 350 pyramidal neurons in a 0.1-0.9 mm range. Excitatory postsynaptic potentials were modeled using a double-exponential function (τ_rise_ = 2 ms, τ_decay_ = 5 ms) and peak conductance of 1 nS. Synaptic strengths *w* decreased exponentially with the distance measured between pre and postsynaptic neurons. Synaptic connections were generated using the *rvolumeconnect* and the associated *rvolumeweight* and *rvolumedelay* functions in parallel GENESIS, quantifying the strength and conduction delay of synaptic contacts between pre and postsynaptic neurons (Bower and Beeman 1998).

The extracellular field potentials recorded by 32 MEA electrodes were simulated in the network model as the sum of transmembrane (including synaptic and ion channel) and capacitive currents from all neuronal compartments of all neurons in the network (n = 156 x 416 x 416) with the help of the 32 GENESIS *efield* objects. The *efield* object uses the electrical conductivity of the surrounding medium to compute the field potential generated by each neuronal compartment based on the pre-calculated distance between a compartment and the *efield* object (i.e. the “virtual” electrode). The field potentials at the “virtual” 32 MEA electrodes were calculated at locations matching the positions of the electrodes in the MEA layout grid using the following equation (Nunez 1981; Nunez and Srinivasan 2006):

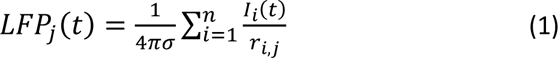

where *I_i_(t)* represents the individual compartment’s current, *r_i,j_*, is the distance from the *i*th compartment to the *j*th MEA electrode contact, and σ = 0.26 S/m is the conductivity of gray matter. The medium in which the neurons were embedded was treated as homogenous and isotropic without capacitive effects. In the case where all MEA electrodes were positioned inside the GM this method of LFP calculation yields potential values that do not differ from the values derived from the reciprocity-based solution for LFP calculation using the FEM (Moffitt and McIntyre 2005). We chose to simulate extracellular field potentials from the network model rather than derive them from the FEM using a reciprocal solution to demonstrate that the recorded/simulated responses reflect predominantly synaptic currents flowing through dendritic arbors of pyramidal neurons. The simulated field potential signals were down-sampled to 30 kHz to match the sampling rate of ground truth data.

### Neural stimulation and recordings

A study participant with spinal cord injury (C5/C6, American Spinal Injury Association (ASIA) Impairment Scale Grade B tetraplegia) was implanted with a total of six MEAs in both hemispheres of the brain (four in left and two in right). Two MEAs (32-channel, 4 mm x 2.4 mm) were implanted in the primary somatosensory cortex (putative Brodmann’s area 1) in the left hemisphere in areas associated with finger and fingertip sensations (McMullen *et al* 2021, Fifer *et al* 2022). The study was conducted under an Investigational Device Exemption (170010) by the Food and Drug Administration (FDA) for the purpose of evaluating bilateral sensory and motor capabilities of microelectrode array implants. The study protocol is registered as a clinical trial (NCT03161067) and was approved by the FDA, the Johns Hopkins Institutional Review Board, and the U.S. Army Medical Research and Development Command Human Research Protection Office.

In previous human studies, vibrotactile stimulations were delivered to fingers where they activated cutaneous mechanoreceptors (McMullen *et al* 2021). The somatosensory cortex responses in areas mapped to tactile receptive field on fingers were recorded and classified as either fast responding (on-off type responses) or slowly responding (unpublished work). In this study, for the purpose of the reconstruction of functional connection in the model we selected a subset of four fast responding (on-off type) electrodes based on their neurophysiological responses to vibratory stimuli. The targeted channels mapped to tactile sensations in the right hand (dominant) on the distal region of the thumb. Pre-selected channels were stimulated individually and ICMS was delivered using a Cerestim R96 (Blackrock Neurotech, Salt Lake City, UT). The ICMS consisted of a single cathodic-first 500 µs charge balanced biphasic pulses (200 µs per phase with 100 µs interphase delay, 80 µA amplitude) delivered at 5 s intervals individually to each targeted electrode. At the same time, we recorded cortical responses from all remaining 31 electrodes. The signals recorded by MEA were buffered (Blackrock Headstage) and amplified (Blackrock Front End Amplifier) before digitizing and storing for offline analysis at 30,000 samples/s. No further digital processing was applied during recording. Recorded cortical responses across 100 trials were aligned to the stimulus onset and averaged for each channel before offline analysis. We also stimulated 4 targeted electrodes at the same time and recorded cortical responses from the 28 remaining electrodes. These recordings were used later to determine whether the cortical model could be “calibrated” to generate the known responses to ICMS delivered to individual MEA electrodes and to predict the cortical response to a superposition of ICMS delivered to 4 electrodes simultaneously.

### Stimulation artifact reduction

After the ICMS presentation, the signal recorded at MEA electrodes is contaminated by a stimulus artifact whose amplitude increased with the number of delivered ICMS pulses. This artifact typically contaminates the recorded cortical responses for several milliseconds (Hao et al 2016, Sombeck et al 2022). To reduce contamination of the recorded signals by this artifact, in this work, we used custom-made patient cables (Blackrock Neurotech) with insulated bundle wires which allowed us to connect the stimulator to the targeted channels and pass signals from the remaining (non-stimulated) channels to the Neural Signal Processor (Blackrock Neurotech). This solution does not eliminate signal saturation on recorded (non-stimulated) channels but reduces the recorded signals contamination by ICMS-induced stim artifact (Fig 7).

### Quantification of the spatiotemporal activity pattern in the somatosensory cortex

During tactile perception, the implanted MEA can record spatial and event-related changes in neural activity in cortical microdomains (cortical micro-volumes surrounding MEA probe tips) mapped to tactile receptive fields on fingers. The neural activity is sampled within a rectangular plane (2.4 x 4 mm) of an implanted MEA with a high temporal (30 kHz) but lower spatial (32 electrodes) resolution. The recorded neural signals reflect cortical processing during tactile perception in a cortical area spanning several adjacent cortical microdomains of Brodmann’s area 1. Identification of functional connectivity between these cortical microdomains requires methods to examine dependencies between field potentials recorded by MEA probe tips in these microdomains after ICMS presentations. Current Source Density (CSD) analysis has been used previously to estimate the location and distribution of synaptic currents based on recordings from multielectrode arrays (Nicholson and Llinas 1975, Colgin 2006, Zhao *et al* 2009, Hindriks *et al* 2017). Here we used a 2D CSD analysis to investigate spatiotemporal activity patterns evoked during cortical responses to ICMS in the MEA plane. CSDs were reconstructed using the kCSD algorithm method (Potworowski *et al* 2012, Chintaluri *et al* 2019). The kCSD is a generalization of the CSD reconstruction method developed for a traditional laminar multielectrode probe (Nicholson and Freeman 75). It allows estimating CSD that generates the potentials measured by electrodes in the case of a planar or 3D high-density electrode arrays. The 2D CSD analysis was performed for both the modeled and recorded extracellular signals and the obtained CSD profiles were used to evaluate the model prediction accuracy. We used (adapted for the CSD) digital image-analysis techniques (Jaleel *et al* 2021) to compare and assess CSDs obtained from model-simulated data with CSDs obtained from recorded (ground truth) data from the study participant. Specifically, we used the thresholding technique (a type of CSD profile segmentation) for CSD comparison which converts a CSD profile into a binary profile and makes the CSD comparison process easy.

### Functional connectivity reconstruction in the computational model of the somatosensory cortex

The reconstruction process for the functional lateral connections in Broadmann’s area 1 of the somatosensory cortex studied required the creation of horizontal synaptic connections in the cortical network model between presynaptic neurons activated in the surroundings of the stimulating electrode and postsynaptic neurons located in the distant surroundings of the electrode where the relevant responses after simulation were detected (Fig 3). These connections may represent the horizontal patchy cortical projections within layer 2/3 (Lund *et al* 1993, Burton and Fabri 1995). The reconstruction process relies solely on field potentials of ground truth data recordings from the study participant (mainly on its low-frequency component). The low-frequency component of MEA recordings can reveal information about the dendritic processing of synaptic inputs that takes place in a larger (i.e. statistically significant) number of neurons (Pettersen et al. 2012; Buzsáki et al. 2012; Einevoll et al. 2013a) as opposed to the high-frequency component which only contains information about the spiking activity of individual neurons near the recording electrodes. In addition, the recorded field potentials represent a deterministic component of cortical responses in contrast to the single or multiunit activity, which are probabilistic (Hao *et al* 2016). Therefore, in this work, the reconstruction relies on information the recorded field potentials reveal to inform the model synaptic connectivity parameters. We assumed that the obtained ground truth recordings encompass a sufficient amount of information to reveal all relevant synaptic connectivity properties in the cortical region of interest necessary for ICMS delivery improvements to elicit more intuitive and meaningful tactile percepts.

**Fig 3.**
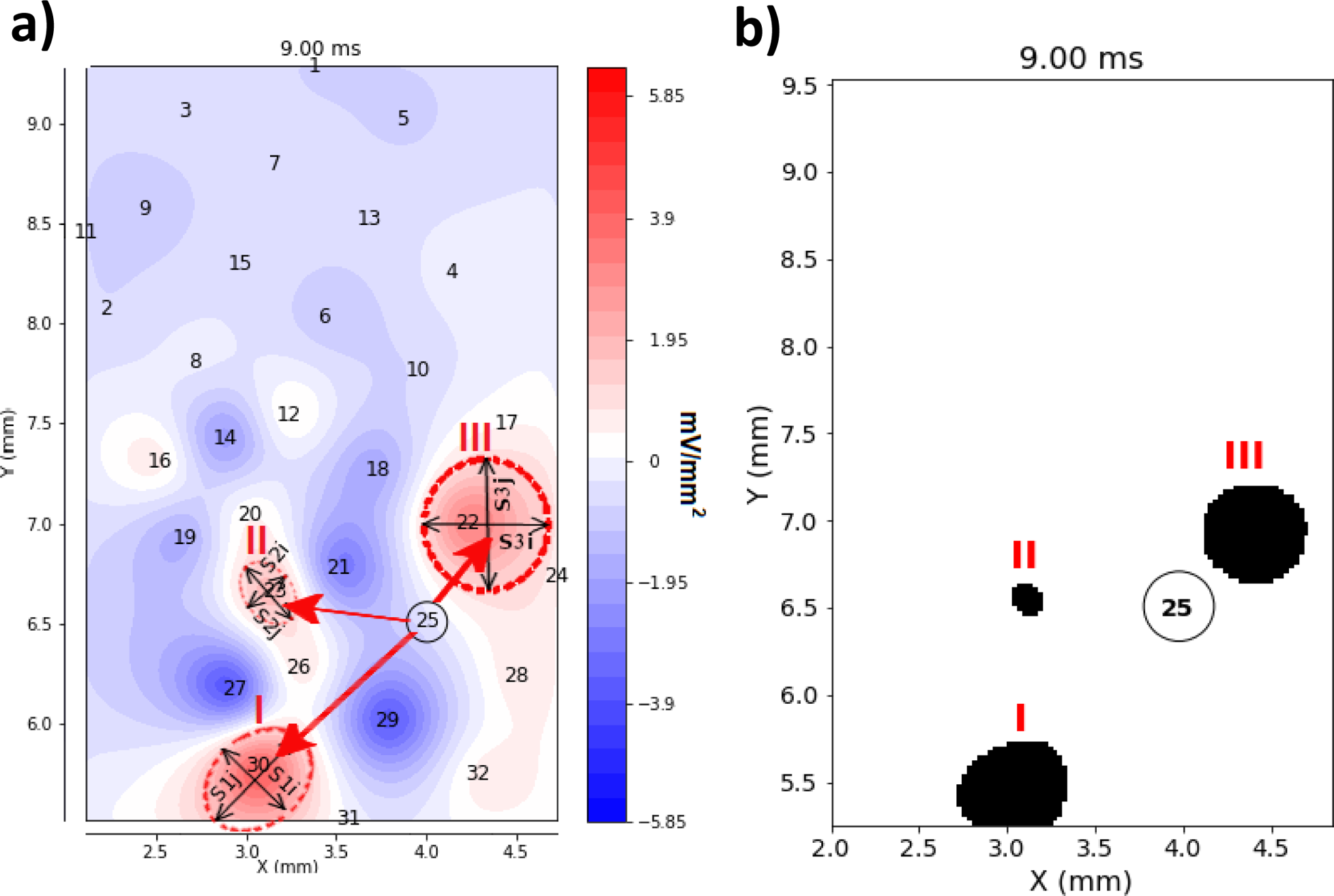
a) The 2D CSD profile was calculated 9 ms after ICMS presentation at MEA contact 25. Current sources I-III (before thresholding) are shown in red, and stimulating MEA contact 25 is circled in black. Current sources (elliptical dashed line) were quantified in terms of the length of the major (S1j, S2j, S3j)/minor (S1i, S2i, S3i) axis and centroid coordinates (mass centers). b) The 2D CSD profile binarized with a threshold > 2 mV/mm^2^. Current sources are labeled with Roman numerals. The computed current source features (centroids and areas) are reported in Tabale 1, Electrode 25 column.

The 2D CSD analyses reveal locations of current sinks and sources in the MEA plane and were used to: 1) assess the pattern of spatiotemporal synaptic activations underlying cortical processing at somatic and dendritic levels of cortical neurons in the surrounding of a recording electrode, and 2) determine the directions of intracortical information flow via the cortical horizontal projections within the area of the somatosensory cortex sampled by MEA electrodes.

The MEA with 1 mm long electrode shanks used in these studies record neural signals at the border of the supragranular and granular cortical layers. We assumed the recorded signal from a single electrode reflects predominantly currents flowing through the basal dendrites of pyramidal neurons associated with neuronal synaptic activation after stimulation. The 2D CSD analyses performed on all neural responses recorded by the MEA allowed the determination of spatiotemporal changes in the distribution of current sinks and/or sources at the basal dendrites of pyramidal neurons within the MEA plane. Current sinks and/or sources were then quantified in terms of shape, area, mass center, perimeter length (features) and used to guide the reconstruction process of the cortical connectivity in the model.

In this work we performed the reconstruction of the functional connectivity based solely on an analysis of current sources because the obtained 2D CSD consisted predominantly of stable current sources that were present 10 - 35 ms after ICMS presentations. These current sources corresponded to vertex-positive deflections recorded typically in 3 - 5 cortical microdomains surrounding the stimulating electrode (Fig 4 Recorded). They indicate hyperpolarization of basal dendrites of pyramidal neurons that “mirrored” current sinks created at the apical dendrites when neurons were synaptically activated via horizontal cortical projections after ICMS application. For each current source we calculated the mass center coordinates (centroids) and the total current source area. We used the obtained values to inform the range and extent of synaptic projection parameters in the model. The sources of synaptic projections were determined based on the area of recruited presynaptic neurons individually for each stimulating electrode (by counting neuronal recruitment in the model). The horizontal connections were created in the model using the *rvolumeconnect* function available in the pGenesis simulator. These connections were symbolically illustrated in Fig 3 by red arrows.

**Fig 4.**
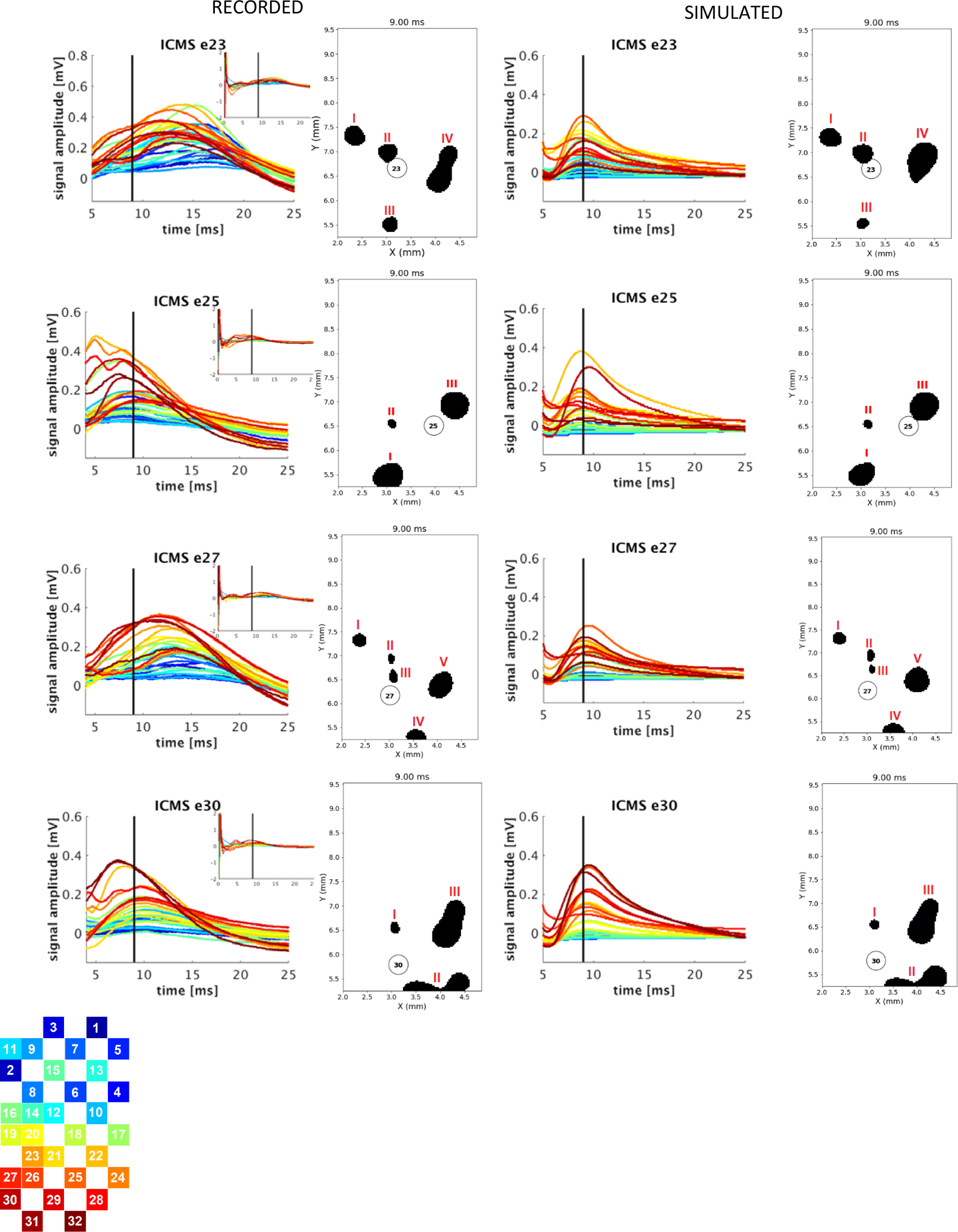
The comparison of recorded (ground truth) and model-simulated cortical responses to a single ICMS pulse applied individually to MEA contacts 23, 25, 27, and 30. The 2D CSD profiles calculated at 9 ms after ICSM presentations (indicated by black vertical lines) are shown next to recorded/simulated neural signals. CSD profiles were binarized with a threshold > 2 mV/mm_2_. Current sources are labeled with Roman numerals. The computed current source features (centroids and areas) are reported in Table 1. The post-stimulation times (> 5 ms) of recorded signals were plotted for better separation of signals recorded by individual electrodes. The position of the color-coded electrode layout on 2D CSD profiles is slightly rotated CCW (approximately by 10 degrees).

**Table 1.** Current source features (centroid coordinates, areas) calculated from the 2D CSD of recorded neural signals (ground truth) and simulated (model predicted) cortical responses to ICMS pulses delivered individually (first four columns) and simultaneously (last column) to microelectrodes 23, 25, 27, and 30. Percentage error shown in column err(%) is a measurement of the discrepancy (absolute value) between the model predicted and the ground truth feature value.

| Electrode #<br>CSD Source # |  | 23 |  |  | 25 |  |  | 27 |  |  | 30 |  |  | 23+25+27+30 |  |  |
| --- | --- | --- | --- | --- | --- | --- | --- | --- | --- | --- | --- | --- | --- | --- | --- | --- |
|  |  | recorded | model | err(%) | recorded | model | err(%) | recorded | model | err(%) | recorded | model | err(%) | recorded | model | err(%) |
| I | Centroid x,y [mm] | 2.54<br>7.34 | 2.55<br>7.33 | 0.4<br>0.1 | 3.11<br>5.71 | 3.07<br>5.73 | 1.29<br>0.35 | 2.56<br>7.34 | 2.57<br>7.33 | 0.4<br>0.14 | 3.13<br>6.65 | 3.15<br>6.66 | 0.15<br>0.63 | 2.55<br>7.48 | 2.53<br>7.34 | 0.78<br>1.87 |
|  | Area [mm<sup>2</sup>] | 0.1 | 0.097 | 3.0 | 0.183 | 0.188 | 2.73 | 0.0465 | 0.045 | 3.23 | 0.027 | 0.023 | 14.81 | 0.227 | 0.164 | 27.75 |
| II | Centroid x,y [mm] | 3.1<br>7.04 | 3.11<br>7.04 | 0.32<br>0 | 3.17<br>3.65 | 3.19<br>6.66 | 0.6<br>0.3 | 3.11<br>6.99 | 3.12<br>7.01 | 1.53<br>1.14 | 3.91<br>5.62 | 4.11<br>5.67 | 5.11<br>0.89 | 3.12<br>7.03 | 3.16<br>6.99 | 1.28<br>0.57 |
|  | Area [mm<sup>2</sup>] | 0.083 | 0.094 | 13.25 | 0.019 | 0.02 | 5 | 0.017 | 0.018 | 5.88 | 0.193 | 0.204 | 5.7 | 0.151 | 0.188 | 24.5 |
| III | Centroid x,y [mm] | 3.14<br>5.74 | 3.1<br>5.76 | 1.27<br>0.34 | 4.23<br>7.0 | 4.14<br>6.98 | 2.13<br>0.29 | 3.15<br>6.67 | 3.14<br>6.69 | 0.32<br>0.29 | 4.05<br>6.70 | 4.06<br>6.77 | 0.25<br>1.04 | 3.42<br>5.67 | 3.65<br>5.62 | 6.72<br>0.88 |
|  | Area [mm<sup>2</sup>] | 0.058 | 0.06 | 3.45 | 0.194 | 0.201 | 3.61 | 0.028 | 0.024 | 14.28 | 0.322 | 0.311 | 3.42 | 0.292 | 0.228 | 21.91 |
| IV | Centroid x,y [mm] | 4.03<br>6.71 | 4.12<br>6.96 | 2.23<br>3.73 | - | - | - | 3.54<br>5.59 | 3.55<br>5.57 | 0.28<br>0.36 | - | - | - | 4.20<br>7.06 | 4.04<br>6.89 | 3.81<br>2.41 |
|  | Area [mm<sup>2</sup>] | 0.255 | 0.262 | 2.67 | - | - | - | 0.0514 | 0.056 | 8.95 | - | - | - | 0.396 | 0.433 | 9.34 |
| V | Centroid X,y[mm] | - | - | - | - | - | - | 3.96<br>6.51 | 3.98<br>6.52 | 0.5<br>0.15 | - | - | - | - | - | - |
|  | Area [mm<sup>2</sup>] | - | - | - | - | - | - | 0.158 | 0.170 | 7.59 | - | - | - | - | - | - |

## Results

### Recorded cortical responses to ICMS

The cortical responses to single ICMS pulses delivered individually to the pre-selected four electrodes (23, 25, 27, and 30) consisted of positive signal deflections that peaked 10 - 15 ms post stimuli that typically returned to baseline 35 ms after the ICMS onset (Fig 4 Recorded ICMS e23, e25, e27, e30). They were present in recordings from electrodes sampling cortical microdomains surrounding the stimulating electrode. Electrodes sampling distal cortical microdomains more than 2 mm away from the stimulating electrode recorded responses of significantly smaller amplitude or did not detect responses at all. Stimulating electrodes were excluded from analyses due to the non-stationary nature of the stimulus artifacts superimposed on the recorded local neural activity. These artifacts had a slowly decaying tail (a negative component) whose amplitude increased with the number of delivered ICMS pulses. In all remaining electrodes, stimulation artifacts consisted of a short ramp-up and down epoch followed by a brief transient period that lasted up to 2 ms before the signal returned to baseline (Fig 4 Recorded ICMS 30 inset). The cortical responses to ICMS pulses appeared consistently in each trial and varied depending on the electrode selected for stimulation. Because observed responses occurred approximately 10 - 20 ms after the ICMS, these responses were unlikely a result of stimulus artifact, which subsides within 2-3 ms after ICMS presentation. Both the range of latencies and the observed between-electrode variability of responses suggest the existence of distinct functional lateral connections across cortical microdomains in Brodmann’s area 1 of somatosensory cortex at the mm scale.

### 2D CSD analyses of cortical responses to ICMS

To compare cortical responses to ICMS pulses delivered to different electrodes, we collected 2D CSD profiles for each stimulating electrode up to 35 ms after ICMS presentation. CSDs were calculated in a 2.4 x 4 mm plane of the MEA (parallel to horizontal cortical connections) at selected time points (every 0.033 ms). Rapidly changing patterns of current sinks and sources that grew and dissipated were found in CSD profiles immediately after ICMS application, while steady and longer-lasting current sources were present later 10 - 30 ms after ICMS pulses. This observation agreed with vertex-positive deflections of cortical responses to ICMS pulses recorded by electrodes in cortical microdomains surrounding the stimulating electrode in Fig 4 (Recorded ICMS e23, e25, e27, e30). We applied thresholding techniques to analyze these steady current sources. A CSD thresholding allows selecting for comparison only the largest and most significant current sources in the cortical response to ICMS pulses delivered to different electrodes. The CSD threshold was set at 2 mV/mm^2^ which corresponded to 20 - 30% of the amplitude of a typically observed maximal CSD value. Distinctive current sources (i.e. different positions, amplitudes, and phases) were identified in CSD profiles for each stimulating electrode (Fig 4 Recorded). The calculated mass centers (centroids) and areas of these current sources for each stimulating electrode are reported in Table 1. These results show that each stimulating electrode elicits a specific and unique pattern of cortical activation and suggests that the ICMS effect is specific to a local functional connectivity exciting laterally across cortical microdomains.

### Model simulations: direct neuronal recruitment by ICMS

Model simulation results demonstrated that ICMS pulses activate predominantly proximal axonal compartments of pyramidal neurons surrounding the stimulating electrode tip. For the applied 85 μA current amplitude of the cathodic-leading ICMS pulse, the lateral extent (measured from the electrode tip axis) of the recruited neurons was typically less than the MEA electrode pitch (400 μm). Fig 5 a, b, and c show three different extents of activation for recruited pyramidal neurons in the simulated cortical layer 2/3 for the three positions of the electrode tip relative to the positions of neuronal somatic compartments (below, middle, and above). Vertical locations of a 50 μm long and 60 μm base-wide electrode tip relative to the somatic compartment layer are shown in the figure insets. Action potentials were initiated predominantly (80 - 95%) in axonal regions proximal to the cell body (first three axonal segments). Action potential initiation in axonal branches occurred in a small fraction of neurons when the electrode tip was positioned above the somatic layer. The electrode tip position above the somatic neuronal compartment increased the chances of axonal branches passing near the electrode tip. The vertical position and inclination of the electrode tip with respect to the laminar arrangement of neurons in the simulated cortical layer affected the number and the lateral extent of neuronal recruitment. The neuronal recruitment by ICMS was most effective when the electrode shank was oriented vertically to the cortical layer and the tip position overlapped with the axon initial segment positions of neurons (Fig 5a, bottom position). The smaller the vertical distance between the base of the electrode tip and the axon initial segments of neurons in the cortical layer, the larger the lateral extent of neuronal recruitment. Cathodic-leading pulses were more effective than anodic-leading pulses activating 75 - 100 % more neurons within a wider lateral extent.

**Fig 5.**
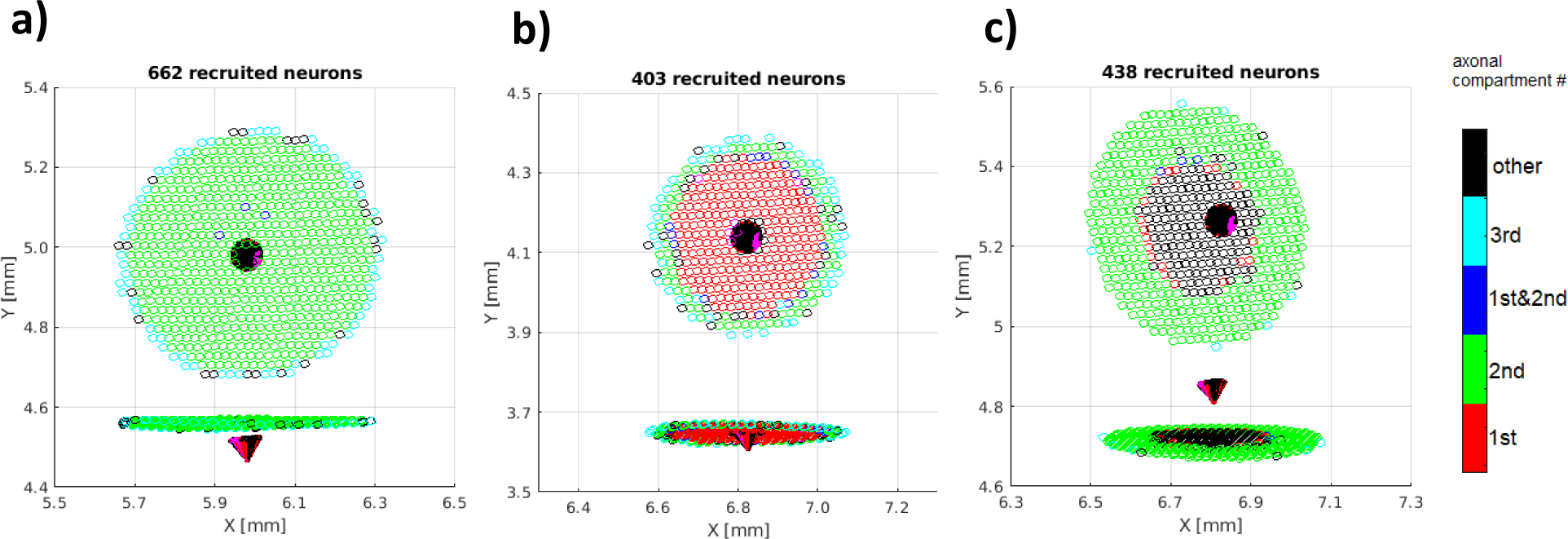
The later extent of pyramidal neuron recruitment in the simulated supragranular cortical layer after ICMS application. Cartesian coordinates (x,y,z) of the soma compartments define the neuronal positions on the coordinate planes (indicated as points), while colors indicate sites of direct axonal activation: red - 1st, green - 2nd, blue - 1^st^ and 2nd, cyan - 3rd axonal compartments (proximal to the cell body). Vertical locations of the electrode tip (red cones) relative to the somatic neuronal compartments are shown in the figure insets: below a), middle b), and above c). The numbers of recruited neurons were 662, 403, and 438, respectively.

### Simulated cortical responses to ICMS

Cortical responses to single ICMS pulses were simulated in the participant-specific model (Brodmann’s area 1) of the somatosensory cortex. Extracellular potentials were computed at locations matching the positions of the electrodes in the MEA layout grid. In the vertical direction, extracellular potentials were simulated at depths equal to the length of the MEA probe shank used in these studies (i.e., 1 mm below the pial surface). This length of the probe shank allowed the recording of neural signals at the border of supragranular and granular cortical layers. Neural signals recorded with 1 mm probe shanks reflected predominantly currents flowing through basal dendrites of pyramidal neurons. The detected short-latency (10 - 20 ms) current sources at the basal dendrites indicated depolarization (current sinks) at apical dendrites when pyramidal neurons were synaptically activated via horizontal cortical projections after ICMS application. The results of the model simulations confirmed that the observed waveforms (positive deflection) in cortical responses were elicited by ICMS and reflected passive current sources at the basal dendrites of pyramidal neurons. These simulated current sources “mirrored” sinks created at the apical dendrites of pyramidal neurons after the arrival of the excitatory synaptic inputs via the simulated lateral cortical synaptic connections activated by the ICMS. Simulations of extracellular potentials were performed in the model after reconstructing functional lateral connectivity between cortical microdomains based on 2D CSD analysis of signals recorded by the 32 MEA contacts. The positive deflections in the simulated extracellular signals were shown in Fig 4 (Simulated).

**Fig 6.**
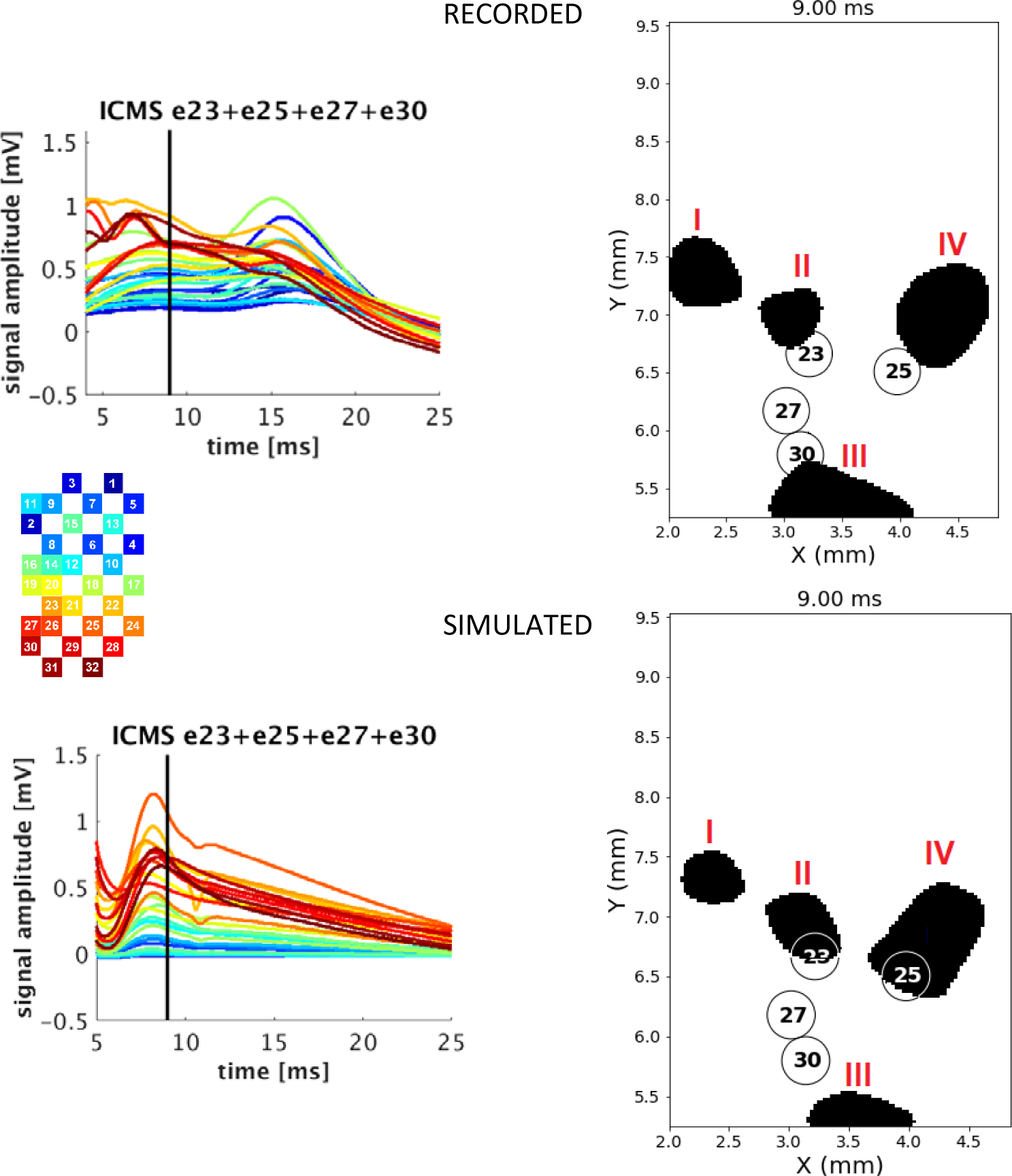
Comparison of the recorded (ground truth) and model-simulated cortical responses to ICMSs applied simultaneously to contacts 23, 25, 27, and 30. The 2D CSD profiles calculated at 9 ms after ICSM application and indicated by black vertical lines are shown on the right next to recorded/simulated signals. CSD profiles were binarized with a threshold > 2 mV/mm^2^. Current source features (centroids and areas) are reported in Table 1. Only the post-stimulation times (> 5 ms) of recorded signals were plotted for better separation of signals recorded by individual electrodes.

### 2D CSD analyses of model simulated cortical responses to ICMS

Reconstructions of functional lateral connectivity between cortical microdomains in the model were performed independently for each stimulating electrode. Current source features (centroid coordinates and area) extracted from 2D CSD profiles of MEA recordings from the study participant (ground truth data) were used to inform the cortical model connectivity parameters and to choose the optimal range and size of patch connections in the cortical model. The range and area of horizontal cortical projections between cortical microdomains have been set independently for each current source identified in 2D CSD profiles in ground truth data recordings for a given stimulating electrode. After connectivity reconstruction, we simulated cortical responses to ICMS delivered to the corresponding electrodes in the model. We performed analogous 2D CSD analyses of the model-derived cortical responses, except that the simulated extracellular signals were multiplied by a factor of 5 to match the range of amplitudes of the signals recorded in the study participant (this factor reflects significantly lower cortical neuron density in the model). As in the case of the ground truth recordings, CSD profiles were calculated for each stimulating electrode, thresholding analyses were applied with a CSD threshold set at 2 mV/mm^2^, and identified current sources were quantified in terms of mass centers (centroids) and area. The comparison of the model simulated current source features to the features of the corresponding current sources calculated from the ground truth data recordings for electrodes 23,25,27, and 30 can be found in Table 1. This method of connectivity reconstruction allowed recreating the 2D CSD profile from the simulated signals with a satisfactory precision for each stimulating electrode. The percentage error calculated as the discrepancy between the model-derived value of a given CSD feature and the ground truth value for the experimental CDS features varied between 0.14 - 5.11% for centroid coordinates and between 2.67 - 14.81% for the calculation of the current source area (Table 1).

### Simulated cortical responses to superposition of ICMS

We reconstructed the functional connectivity in the model to align the 2D CSD profile of the simulated cortical responses to the corresponding CSD profile of the recorded cortical responses for each stimulating electrode. To validate and test the model’s performance, we simulated cortical responses to ICMS pulses delivered to all four electrodes simultaneously. We compared the model-generated cortical responses to the responses recorded from the study participant after ICMS delivery to all four electrodes. We found that the model captures the cortical responses well on a fine timescale as assessed by calculating the values of the 2D CSD features taken between the predicted and recorded (ground truth) responses. The comparison of the 2D CSD features predicted by the model to the 2D CSD features calculated from the ground truth data **is** shown in Table 1 (last column). The model predicted the 2D CDS features of the cortical responses to ICMS delivered to all four electrodes with the percentage error of the centroid coordinate prediction ranging between 0.57 - 6.72% and 10 - 28% for the current source area prediction.

**Fig 7.**
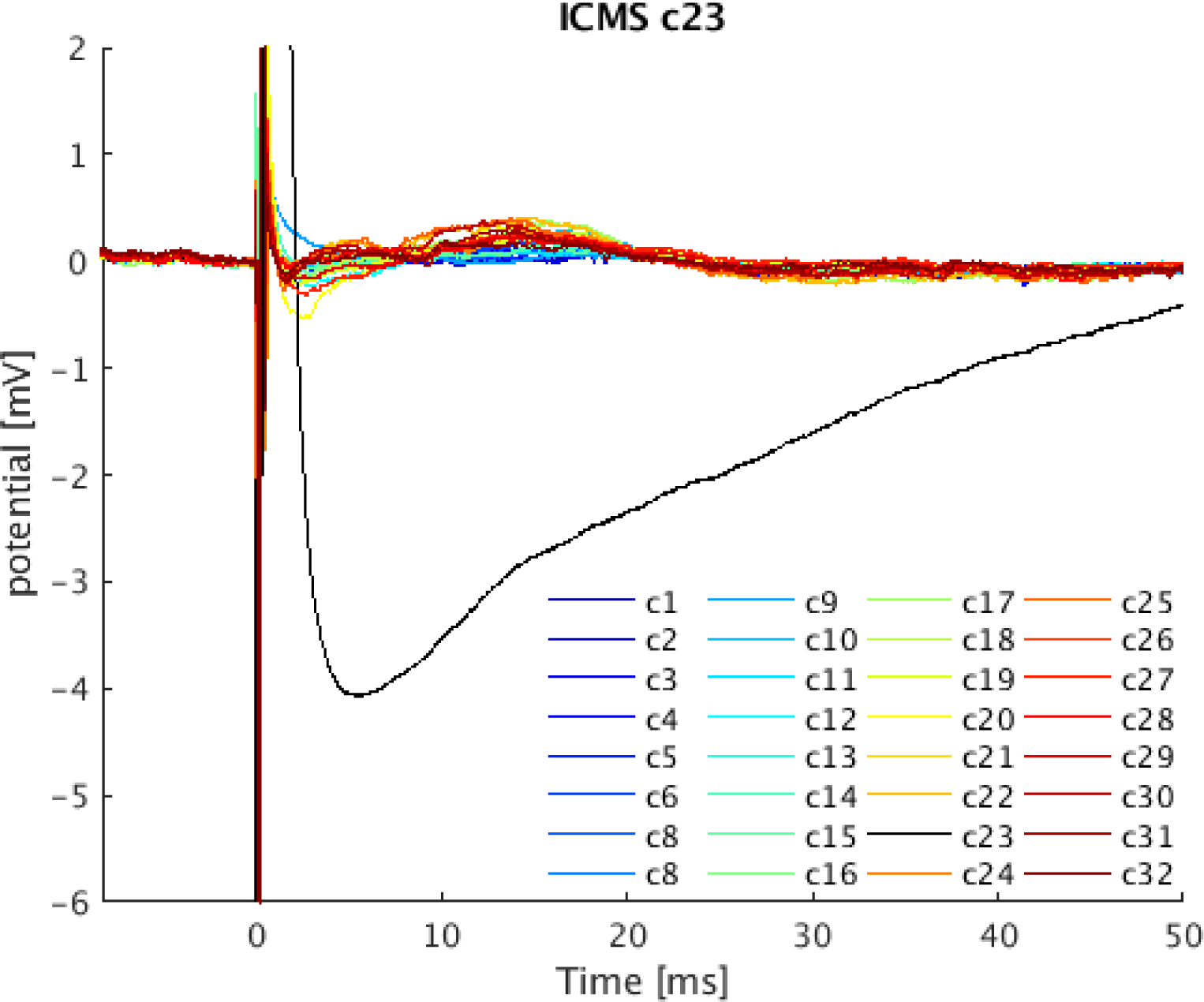
The cortical responses recorded by MEA electrodes after a single ICMS pulse applied to electrode 23. We used custom-made patient cables (Blackrock Neurotech) with insulated bundle wires which allowed us to connect the stimulator to the targeted channels and pass signals from the remaining (non-stimulated) channels to the Neural Signal Processor (Blackrock Neurotech). This solution does not eliminate signal saturation on recorded (non-stimulated) channels but reduces contamination of the recorded signals by the stimulation artifact. For comparison, the ICMS-induced artifact on stimulating electrode 23 (black) has been included.

## Discussion

Optimization of tactile feedback via ICMS in prosthetic devices using only psychometric tasks is extremely time-consuming (Bensmaia 2015) and unlikely to yield biomimetic stimulation patterns. In this work, we presented a model-based approach to predicting cortical responses to ICMS-, as an initial step towards model-optimized ICMS parameter selection. It has been recently shown that a computer model-based approach can aid in designing ICMS patterns needed to evoke naturalistic patterns of neuronal activation in the somatosensory cortex (Kumaravelu *et al* 2022). Our model differs from this modeling study in several ways. Most importantly, Kumaravelu and colleagues attempted to deliver ICMS in a way that recreates the spike pattern of cortical neurons recorded in the somatosensory cortex, while in our model, we focused on extracellular signals recorded by MEA (local field potentials). We built the model of the segment of the human somatosensory cortex implanted with MEA based on the participant-specific 3-D anatomical data (MRI). We then tuned this cortical model to produce simulated cortical responses (field potentials) that matched the cortical responses to ICMS recorded from this study participant. It must be emphasized that in this work, we did not aim to deliver an overfit model but to test the ability of a high-resolution cortical model to predict cortical responses to ICMS delivered to individual and multiple electrodes. We demonstrated that a high-resolution cortical network model, if calibrated to produce the known cortical responses to single ICMS pulses delivered to individual electrodes, could predict with a reasonable accuracy the cortical response to ICMS pulses delivered simultaneously to multiple electrodes. These preliminary results, however, suggest that a high-resolution computer model can potentially predict responses to any arbitrary repeating pattern of ICMS pulses, and therefore it could be useful in selecting the optimal stimulation pattern for restoring somatosensation. The presented ICMS model utilizing anatomically and electrically accurate volume conductor models and multicompartment representations of cortical neurons represents an advanced tool for simulating the effects of ICMS. Such high-resolution cortical models can provide a more comprehensive assessment of the magnitude and extent of neural activity induced by ICMS than has been possible to obtain using extracellular recordings. Identifying the morphology and location of cells activated by ICMS in a cortical model allows us to make predictions about the effects of ICMS *in situ* in neural tissue.

Despite the complexity of this ICMS model, there were several simplifications and assumptions introduced to this model to reduce the burden of implementation, make the numerical simulation tractable, and the modeling tasks achievable. Foremost among these simplifications is the assumption that layer 2/3 pyramidal neurons were the main contributors to the recorded neural signals. A simulation of a large number of biologically-realistic and independent extracellular signals required a reasonable number of multicompartmental neurons with realistic numbers of synaptic connections (spanning across computing nodes) per each simulated extracellular signal. As the maximal number of neurons and synapses per computing node was fixed (and represented a bottleneck for scaling up the model), we focused attention on cortical neurons with the highest contribution to extracellular signals and built the model upon cortical layer 2/3. The entire network model consisted of one cortical layer 2/3 distributed and simulated across 256 computing nodes. An inevitable consequence of the above assumption is that the neuronal synaptic interactions, the main contributors to the simulated neural signals, were reduced in this model to synaptic interactions between pyramidal cells (one neuron type). It is not the case *in situ* where typically neuronal elements of various types of cortical neurons can be activated by ICMS causing various types of synaptic interactions at different time and spatial scales (Osborn *et al* 2021). In our studies, however, we restricted the analysis of recorded neural signals to a short post-stim window (< 30 ms). We hypothesized that early after stimulation, the recorded signals reflect primarily activation of lateral axonal projections and interaction of pyramidal neurons in layer 2/3. A good agreement between the model generated and experimental data justified this assumption. It is important to note that the method of lateral cortical connection reconstruction based on the extracted 2D CSD features does not only apply to the simulated in this model connections between pyramidal neurons in cortical layer 2/3. The 2D CSD features may guide the reconstruction of cortical connectivity between other cortical neurons (if simulated in the model) within or between different cortical layers.

### The role of horizontal (patch) cortical connections

We hypothesized that horizontal (lateral) cortical connections play a central role in mediating tactile sensation. In addition, we hypothesized that these horizontal connections are the main contributor to the early cortical responses to ICMS recorded in the somatosensory cortex. Horizontal connections are a prominent feature of cortical intrinsic circuitry (Gilbert and Wiesel, 1979; Rockland and Lund 1982; Jones *et al* 1979). These connections originate primarily from pyramidal cells, extend for 2–6 mm parallel to the cortical surface, and terminate in a highly selective and patchy manner targeting predominately (80%) excitatory cells (McGuire *et al* 1991). In the striate cortex of primate and cat, neurons targeted by the patchy lateral long-range connections tend to have similar feature preference as their presynaptic origin, resulting in connections linking patches of neurons that have similar orientation preferences (Buzás et al. 2006; Gilbert et al. 1996). Similar horizontal connections are formed by the axon collaterals of pyramidal neurons in the supragranular cortical layer 2/3 of the somatosensory cortex that extend for millimeters parallel to the cortical surface (Lund *et al* 1993, Lund *et al* 2003, Burton and Fabri 1995, Friedman *et al* 2020). In the primary somatosensory cortex, horizontal connections in supragranular cortical layers integrate information across sensory maps by connecting related functional columns. Studies in human and non-human primates have identified areas 3b and 1 in the primary somatosensory cortex as crucial for tactile perception (Bohlhalter *et al* 2002; Kaas 2004). Recent studies in non-human primates have suggested that the patchy system of intrinsic lateral connections in the somatosensory cortex may play an important role in tactile information integration and processing (Négyessy *et al* 2013, Ashaber *et al* 2014). It includes both Brodmann areas 3b and 1, but the role of lateral connections in the latter, which performs the higher-level operations in the hierarchy of somatosensory processing, is still poorly understood. Compared with area 3b, the larger spread of intrinsic lateral connections in Brodmann area 1 could be an anatomical substrate of this higher-level processing during a tactile sensation and percept, which requires and happens at a larger spatial and temporal scale.

### Multiple ICMS temporal patterning for restoring a tactile percept

Considering the results cited in the previous section, efforts to elicit intuitive and meaningful tactile percepts by delivering ICMS to a single electrode implanted in area 1 could potentially be improved by using more biomimetic approaches. Temporal patterning of ICMS has been suggested as a possible approach for encoding complex tactile information and conveying this information more efficiently to the somatosensory cortex (Grill 2018, Eles *et al* 2021). Temporal patterning of ICMS delivered to a selected subset of implanted microelectrodes in an attempt to artificially recreate a typical “natural” pattern of neuronal activation has perhaps the greatest potential and can be the most effective method for restoring intuitive and meaningful sensation. The model-based approach described in this work for restoring the ICMS-evoked sensations is conceptually similar to the “replay stimulation” approach used in studies of the monkey somatosensory system by Weber and colleagues (Weber *et al* 2011). Spatiotemporal neural activity patterns recorded during tactile percepts by 32 MEA electrodes in the somatosensory cortex could serve as templates of which ICMS applied to a subset of microelectrodes should emulate in the cortex. Unfortunately, the effect of ICMS applied to a subset of microelectrodes and delivered to a given tissue volume is very difficult to predict and largely unknown. To our knowledge, no method exists for finding an optimally biomimetic sequence and parameters of individual ICMS for a given spatiotemporal template. Here we showed that a biologically realistic cortical network model can potentially serve as a reliable predictor of cortical response to a given sequence of ICMS presentations. It is important to note that the high-resolution cortical model presented here was developed based on a limited set of neural recordings and the reconstruction of the functional cortical connectivity in this model is not complete. It was performed based on analyses of cortical responses to ICMS delivered to only four stimulating microelectrodes in one study participant. Moreover, it is unclear whether the reconstructed connections in this cortical model are subject-specific or reflect the general connectivity of the human somatosensory cortex. One should remember that the recorded cortical responses are sensitive to the MEA site implantation. We can speculate that the recorded cortical responses may vary between subjects with a sensory loss depending on the type and progression of the injury while may be similar in subjects with normal tactile perception. Because this study was conducted retrospectively and included only one participant, we could not confirm the above speculations and test the ability of this model to make new predictions. We showed, however, that a model with partially reconstructed cortical connectivity was able to predict neuronal responses to ICMS delivered to four microelectrodes simultaneously with considerable accuracy as assessed by the calculated values of the 2D CSD features between the predicted and recorded (ground truth) cortical responses. High-resolution cortical models have been used previously for prediction of the effects of depth (DBS) or subdural cortical stimulation (Gunalan *et al* 2018, Kudela and Anderson 2015, Kudela and Anderson 2021,). Given the predictive power of these models, we hope that the optimal parameters and ICMS orders can be determined based on matching the 2D CSD of simulated extracellular signals from the calibrated model to the 2D CSD of templates recorded by MEA extracellular signals during natural tactile sensations.

## Data Availability

All data produced in the present study are available upon reasonable request to the authors.

## Authorship statement

P.K. performed simulations, interpreted data, wrote manuscript, acted as corresponding author. W.S.A. led implantation planning and execution, reviewed data analysis, and provided revisions to the manuscript. M.S.F., L.E.O. collected data, reviewed data analysis, and provided revisions to the manuscript. D.P.M. contributed to various components of the cortical mapping, experimental design and stimulation testing with implanted arrays. F.V.T. led the clinical study, experimental design and stimulation testing with implanted arrays. G.L.C. clinical study concept and design. P.A.C. led the clinical study; preoperative fMRI planning.

